# Machine Learning-Based Risk Stratification for Gestational Diabetes Management

**DOI:** 10.1101/2022.06.11.22276278

**Authors:** Jenny Yang, David Clifton, Jane Hirst, Foteini Kavvoura, George Farah, Lucy Mackillop, Huiqi Lu

## Abstract

Gestational diabetes mellitus (GDM) is often diagnosed during the last trimester of pregnancy, leaving only a short timeframe for intervention. However, appropriate assessment, management, and treatment have been shown to reduce the complications of GDM. This study introduces a machine learning-based stratification system for identifying patients at risk of exhibiting high blood glucose levels, based on daily blood glucose measurements and electronic health record (EHR) data from GDM patients. We internally trained and validated our model on a cohort of 1,148 pregnancies at Oxford University Hospitals NHS Foundation Trust (OUH), and performed external validation on 709 patients from Royal Berkshire Hospital NHS Foundation Trust (RBH). We trained linear and non-linear tree-based regression models to predict the proportion of high-readings (readings above the UK’s National Institute for Health and Care Excellence [NICE] guideline) a patient may exhibit in upcoming days, and found that XGBoost achieved the highest performance during internal validation (0.021 [CI 0.019-0.023], 0.482 [0.442-0.516], and 0.112 [0.109-0.116], for MSE, R2, MAE, respectively). The model also performed similarly during external validation, suggesting that our method is generalizable across different cohorts of GDM patients.

## 1. Introduction

Gestational diabetes mellitus (GDM) is one of the most common health conditions during pregnancy, with a prevalence of one in six pregnant women worldwide [1]. GDM is defined as glucose intolerance of either first onset or recognition during pregnancy [2]. It is associated with both maternal and fetal complications, including perinatal death, excessive fetal growth (leading to problems during childbirth), preeclampsia, and neonatal hypoglycemia. Additionally, women who develop GDM are at increased risk of developing Type 2 diabetes [3,4,5]. Appropriate assessment, management, and treatment have been shown to reduce the complications of GDM. However, GDM is typically diagnosed during the last trimester of pregnancy, thus leaving only a short timeframe for intervention (typically around 10-16 weeks) [6]. Once diagnosed, counselling is provided for lifestyle management, including dietary and exercise modifications. To monitor improvement, women are asked to check their glucose levels through finger stick-testing several times a day. In the case that glucose levels remain high, medication is prescribed [3].

The UK’s National Institute for Health and Care Excellence (NICE) guidelines states women with GDM require an increased level of maternal and fetal surveillance as they may need more interventions during pregnancy [7]. Based on the NICE guidelines, nurses and clinicians should review women with GDM at least once every two weeks, from the time of diagnosis of GDM until delivery. However, women who are at high risk of hyperglycaemia (high blood sugar level), or those who, despite treatment, demonstrate persistent hyperglycaemia, can require more frequent clinical review. Given that many NHS Trusts and hospitals abroad provide care for very large numbers of women with GDM at any one time (often > 100 women), this presents a challenge for busy clinicians. Traditionally, glucose control has been assessed based on historic blood glucose readings recorded by the women in paper forms or diaries. The advent of digital monitoring opens new possibilities for prediction of women at risk of hyperglycaemia.

In clinical practice, clinicians typically review patients’ blood glucose levels every 2-4 weeks in the outpatient antenatal clinic. Thus, with the advent of real-time monitoring using GDm-Health, there is the potential for more frequent review and more responsive medication adjustment. However, this needs to be balanced against workload generation for the clinical team. We propose a novel data-driven model that can bypass the need to manually screen all patients with high blood glucose readings before making decisions on patient-review order, significantly reducing the burden on clinicians. Specifically, we introduce a model for stratifying patients with GDM based on hyperglycaemia risk, and consequently, their need for clinical review. Using machine learning-based regression models, the proposed pipeline quantifies the risk of hyperglycaemia for the three days immediately following a blood glucose measurement.

## 2. System Design

### 2.1. Bluetooth-enabled Blood Glucose Measurement

To address the challenges of monitoring patients with GDM and providing timely intervention, the University of Oxford and Oxford University Hospitals NHS Foundation Trust (OUH) developed a digital blood glucose management system, GDm-Health [6]. This is a smartphone-based, Bluetooth-controlled blood glucose monitoring system, enabling remote self-monitoring and bidirectional communication between patients and clinicians.

Women who were diagnosed with GDM and managed by OUH were subscribed to the GDm-health system to monitor their blood glucose levels. During the period of using GDm-Health, women were recommended to measure their blood glucose between four to six times a day (recorded as pre-breakfast, one-hour post-breakfast, pre-lunch, one-hour post-lunch, pre-dinner, and one-hour post-dinner), for a minimum of three days of the week [7]. We will refer to these six measurements as “Tags” in this study.

Additionally, GDm-Health has a heuristic alerting system based on clinical care pathways used by OUH during the development of GDm-Health. A red flag is generated if three or more consecutive blood glucose readings are above the designated threshold at the same meal tag. An amber flag is generated if two or more consecutive readings are above the threshold [6]. However, with large numbers of women with GDM managed in each hospital, a significant proportion of women have red or amber flags every week, generating substantial work for clinicians. Thus, there is a need for improved approaches to further stratify patients within these higher risk groups, who need urgent attention.

We developed a model that aims to streamline clinician workflows by automating the identification of patients that need urgent clinical review. This algorithm can be used as an intelligent add-on module on GDm-Health or as a stand-alone system for any GDM clinic if they have access to patients’ daily blood glucose data.

### 2.2. Participant Inclusion and Exclusion

For this study, we used de-identified, linked electronic health record (EHR) data and blood glucose measurements. Women with GDM, managed at the OUH, and subscribed to the GDm-Health system between April 30, 2018 to May 4, 2021 were included in this study (1,148 pregnancy cases). Patients with more than one pregnancy during the study period were considered for each pregnancy, independently. Additionally, we only considered patients who had one baby during the pregnancy (i.e., twin pregnancies were not included in the study). Pregnancies with less than 36 blood glucose readings were excluded to ensure that models were trained on patients who have established their blood glucose test patterns. This threshold of 36 readings assumed that patients would establish their blood glucose test pattern after 7-10 days of blood glucose monitoring (with 4-6 readings per day). Excluding patients with a small number of blood glucose measurements reduces the risk of having distribution shifts between training and prediction samples, removing possible biases, such as patient behavioural changes.

To externally validate the performance of our model, the GDm-Health data of 709 pregnancy cases at the Royal Berkshire Hospital was also included using the same inclusion and exclusion criteria.

### 2.3. Hyperglycaemia Risk Score Definition

The model is designed based on the hypothesis that the predicted three-day mean percentage of high readings, immediately after a three-day observation window, can be used as a proxy for hyperglycaemia risk. This score can then be used to stratify patients in need of clinical review, supporting clinicians in deciding whom to review based on predicted risk.

## 3. Methods

### 3.1. Data Preprocessing

There were 1,148 pregnancies and 272,712 blood glucose readings considered during the model development process. In our study, the measurements used were self-tested and self-reported by patients; thus, the blood glucose testing frequency varied. As shown in Fig. 1, the highest numbers of blood glucose measurements were taken pre- and post-breakfast, post-lunch, and post-dinner (in the OUH cohort).

**Figure 1:**
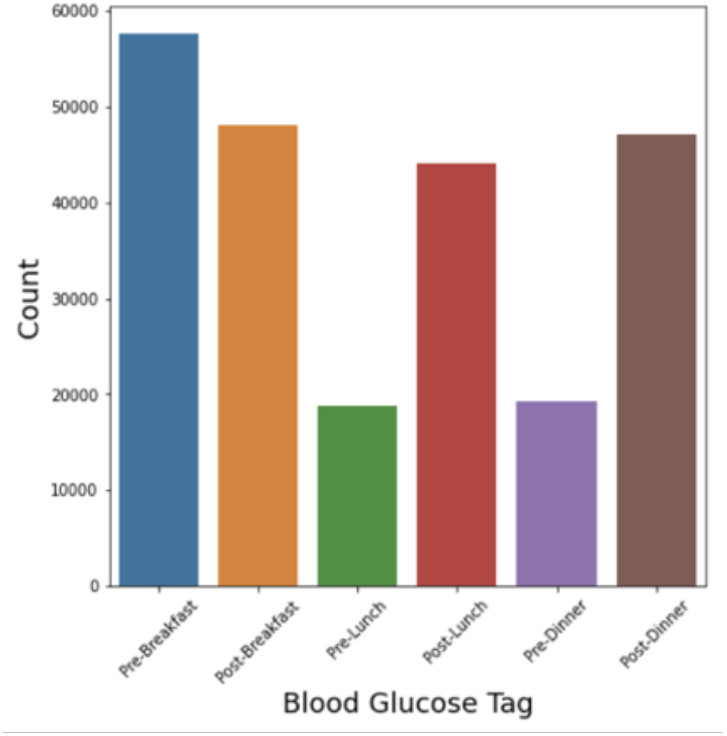
Blood glucose readings for each tag across patients in the OUH cohort. 105,693 readings recorded pre- and post-breakfast, 62,923 recorded pre- and post-lunch, and 66,429 recorded pre- and post-dinner.

**Figure 2:**
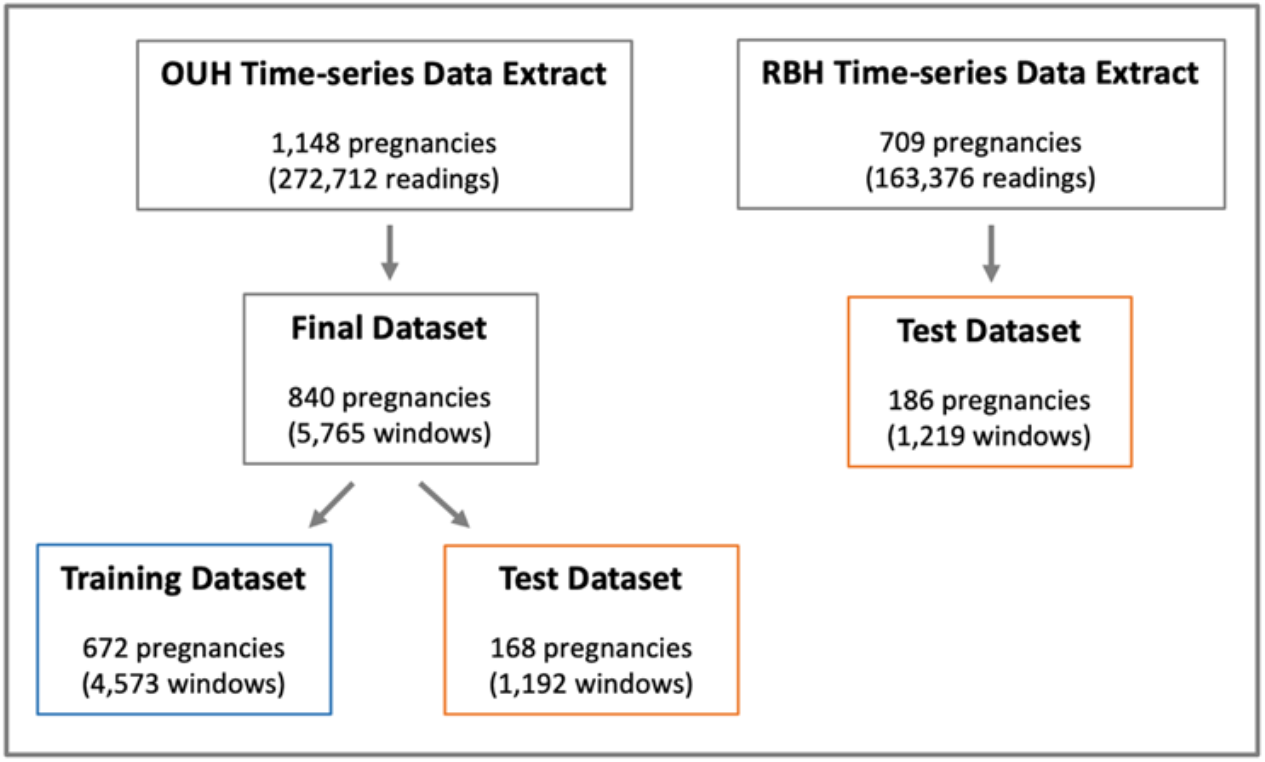
Final datasets after all data exclusion considerations, pre-processing methods, and training/testing split (80:20) of the OUH dataset.

Beyond the difference in sample rate frequencies, the duration of blood glucose monitoring (between the week of recruitment and the week of giving birth), also varied. Thus, for each pregnancy, we separated blood glucose measurements into multiple three-day windows, using measurements from one three-day window to predict the hyperglycaemia risk for the following three-day window. This windowing method helps minimize any possible errors from outliers or missing measurements over the three-day period. Furthermore, it allows us to use more of the data, as days with some missing tag values can still be considered in training. The risk score being predicted (score between 0-1, with 1 representing the highest risk), is defined as the proportion of blood glucose readings above the NICE advised thresholds [7]. A higher proportion of high-reading alerts indicates a higher risk of blood glucose abnormality. This set-up allows for blood glucose status to be predicted after each blood test, making it easier for clinicians to stratify patients with abnormal blood glucose levels. Additionally, as the monitoring period between clinical review for GDM patients is typically between three-days (and possibly up to two weeks), the window size is suitable for current clinical review periods. The ability to prioritize patients with higher hyperglycaemia risk, helps enable personalized and continuous GDM blood glucose monitoring.

In the OUH time-series data, we further removed any readings with miscellaneous values, such as those recorded as NaN or those with invalid time tags (i.e., tags not corresponding to any of the six). We did not filter any extreme blood glucose values outside reasonable ranges, as we wanted to develop a model that would be robust to any errors that may be due to data collection/sensor recordings. Blood glucose readings under the “pre-lunch” and “pre-dinner” tags were also excluded during model development, as these tags had far fewer measurements recorded (less than half of the frequency of the other tags). By removing these, we were able to avoid using any form of data imputation for missing values. Moreover, the NICE guideline advises patients to have four measurements every day; thus, developing a model based on four tag features is well-suited for the task, without overwhelming women with additional blood tests. We then combined the remaining blood glucose readings with their linked, de-identified EHR features. We included three features previously shown to affect blood glucose levels – maternal age, gestational day (duration of pregnancy in days), and medication [8,9,10]. Pregnancies missing any of these features were excluded, leaving 840 pregnancies (collectively contributing 5,765 windows) in the final dataset.

Although BMI has been previously found to affect blood-glucose levels, there were many missing values present in our dataset, and since we removed any windows with missing values, including BMI as a feature would significantly reduce the size of our training set (from 4,573 windows to 2,500). Thus, we did not include it as a feature in our primary analyses.

We applied the same preprocessing pipeline to our external validation dataset collected at the RBH, which resulted in 186 pregnancies (corresponding to 1,219 windows) available for testing (screened from 163,376 blood glucose readings, from 709 patients).

Beyond the time-series blood glucose readings and EHR features, we also generated two engineered features. The first feature is based on high blood glucose readings, which considers any blood glucose measurements higher than the NICE advised blood glucose ranges, which are defined as target measurements between 3.5 mmol/L and 5.8 mmol/L for fasting measurements, and measurements less than 7.8 mmol/L for 1-hour postprandial measurements [7]. Using this definition, we calculated the value of this feature as the percentage of high-readings that occur within the three-day observation period (across all tags). The second engineered feature we calculated is the average rate of change of blood glucose measurements over the three-day observation window (calculated individually for each time tag). We refer to these as High-readings and Gradients, respectively. The full summary of features (and their respective definitions) included in model development are listed in Table 1. We grouped individual features into corresponding sets based on accessibility, namely, sensor-provided features (Tags), two types of engineered features (Gradients, High-readings), and EHR data (maternal age, gestational day, medication).

**Table 1:** Feature sets used in model development.

| Feature Set | Features | Definition |
| --- | --- | --- |
| **Tags** | Pre-breakfast reading, Post-breakfast reading, Post-lunch reading, Post-dinner reading | *Tags* correspond to mean blood glucose measurements for a given time-point (tag), over the three-day observation period |
| **Gradients** | Pre-breakfast gradient, Post-breakfast gradient, Post-lunch gradient, Post-dinner gradient | *Gradients* correspond to rate of change of blood glucose for a given time-point (tag), over the three-day observation period |
| **High-readings** | Percentage of high readings | *High-readings* is the percentage of high-readings among all blood glucose measurements within the three-day observation period. This feature is also calculated for the subsequent three days (three days following the observation window) and used as the predicted output of the models. |
| **EHR** | Maternal age, Gestational day, Medication | *Maternal age* is the age of the woman when she is confirmed with pregnancy.<br>*Gestational day* is the average day of the woman's pregnancy (gestational) days over each three-day window.<br>*Medication* is a binary feature, defined as anyone taking Metformin and insulin during pregnancy. |

Summary population characteristics of OUH and RBH patient cohorts are reported in Table 2, and summary population statistics of the features used in training can be found in Supplementary Tables B1 and B2, for the OUH and RBH cohorts, respectively.

**Table 2:** Patient characteristics and statistics for OUH and RBH cohorts. Mean values recorded alongside standard deviation (where applicable). *missing values present

| Characteristic | OUH Cohort | RBH Cohort |
| --- | --- | --- |
| Age (years) | 33.6 (5.2) | 33.6 (4.9)* |
| BMI | 31.0 (6.8)* | 28.6 (6.9)* |
| Ethnicity | Not Stated: 506 |  |
|  | White: 231 | Not Stated: 43 |
|  | South Asian: 44 | White: 63 |
|  | Black: 27 | South Asian: 66 |
|  | Other: 22 | Black: 7 |
|  | Chinese: 8 | Other: 7 |
|  | Mixed: 2 |  |

### 3.2. Model Development and Hyperparameter Optimization

For hyperglycaemia risk prediction, we trained both linear and non-linear ensemble models. To predict a continuous risk score, we used multiple linear regression (MLR), Random Forest, and XGBoost regression models. All models can handle tabular data consisting of both continuous and categorical features. MLR is a parametric model that is widely accepted in clinical decision-making, making it an appropriate benchmark for comparison to more complex models. Random Forest is an ensemble method built on decision trees, and XGBoost is an optimized distributed gradient boosting library which has been found to outperform Random Forest and other tree-based models. It is an ensemble model that has achieved state-of-the-art results on many machine learning challenges, especially those involving structured or tabular datasets (as we are using in our study). Another benefit of using tree-based models is that feature importance can be explained using Shapley additive explanations (SHAP).

To predict the impending proportion of high-readings a woman will have in the upcoming three days, we used the features in a three-day window as the input features (representing the features, X, Table 1) and the percentage of high-reading alerts in the subsequent three-day window as the output (corresponding label, y, respectively). Each X and y pairing is then treated as an individual sample during model development. To understand the role of each feature set (Tags, Gradients, EHR, High-readings) in MLR, Random Forest, and XGBoost models, we reported model performances using a stepwise method, adding additional feature sets one at a time for each subsequent model developed, thereby evaluating their relative importance.

To choose the appropriate training settings for the XGBoost regressor, we plotted the model outcome variable (i.e., percentage of high-readings alerts over the subsequent three days), y, to look at its distribution. As shown in Fig. 3, this variable is highly skewed, with many zeros. However, as our model focuses on predicting patients at risk of exhibiting high blood glucose levels, we chose to use a Gamma distribution to represent the distribution of the predictor variable, such that our model is focused on non-zero values for high-readings.

**Figure 3:**
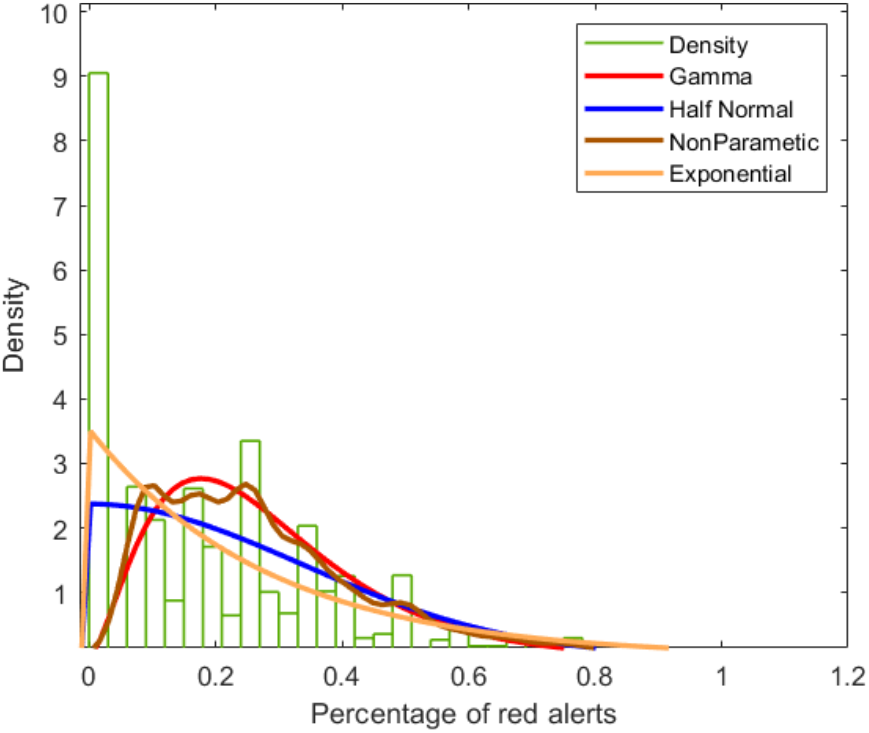
Percentage of high-readings in three-day prediction windows to be predicted (OUH dataset), compared to different distributions.

For model development, we used an 80:20 ratio of the OUH data, by individual pregnancy, resulting in 4,573 and 1,192 windows in the training and test set, respectively (corresponding to 672 and 168 patients, respectively). We used the training set for hyperparameter optimization and modeling training. For the XGBoost model, we implemented a grid search for different values of the learning rate, number of trees used, maximum tree depth, percentage of samples used per tree, and percentage features used per tree. Standard five-fold cross-validation was then applied to evaluate which hyperparameter combination performed the best. We used the same number of trees and maximum tree depth in the Random Forest Model. Details about the final settings and hyperparameter values used for each model can be found in Supplementary Table C1. After successful hyperparameter optimization, we tested the final model on the held-out test set.

We started model training by using only tag features, as a baseline model, and sequentially added additional feature sets. The availability of feature sets can vary depending on the level of data access in different settings; thus, we evaluate all combinations of these sets in model development. Although we did not include BMI in our primary analyses, we tested its influence using the subsequently reduced training set, to demonstrate the potential of including it as a feature in any future analyses. Results for this can be found in Appendix D in the Supplementary Material.

To compare our models, we reported the mean squared error (MSE), the R2 value (R2), and the mean absolute error (MAE). Because our goal was to stratify patients at risk of having high blood glucose levels, the actual prediction value itself was ancillary to the order in which patients are ranked. Thus, we also considered the accuracy in which a patient is ranked. We determined this rank by calculating the percentage of patients that were correctly triaged into correct risk bounds. We considered three label-encoded scoring bounds – lower-, middle-, and upper-bounds (scores binned by equally splitting lower-, middle-, and upper-thirds). To further understand the contribution of individual features to model predictions, we also performed SHAP analysis.

## 4. Results

### 4.1. Model Training and Internal Validation

Both tree-based ensemble methods outperformed MLR (Table 3), and the narrow CIs showed model stability. The XGBoost model achieved the best performance in terms of all metrics (0.021 [CI 0.019-0.023], 0.482 [0.442-0.516], and 0.112 [0.109-0.116], for MSE, R2, MAE, respectively), followed closely by the Random Forest model (0.022 [0.021-0.024], 0.447 [0.400-0.482], and 0.117 [0.117 (0.114-0.121], for MSE, R2, MAE, respectively). The MLR model performed significantly poorer, achieving 0.035 [0.031-0.149], 0.155 [0.000-0.179], and 0.142 [0.127-0.150], for MSE, R2, and MAE, respectively. The XGBoost model also achieved the highest accuracy for all three risk bounds (0.609 [0.582-0.633], 0.413 [0.387-0.438], and 0.650 [0.624-0.675] for lower-, middle-, and upper-bounds, respectively) compared to the other two models; and as before, the Random Forest model performed second best (0.598 [0.569-0.625], 0.404 [0.378-0.430], 0.639 [0.610-0.665] for lower-, middle-, and upper-bounds, respectively). Results between the XGBoost and MLR models were significantly different (Wilcoxon Signed Rank Test, p < 0.0001), as well as between the XGBoost and Random Forest models (p < 0.0001).

**Table 3:** Performances of MLR, Random Forest, and XGBoost models trained on Tags feature set. Results are reported alongside 95% confidence intervals (calculated using 1000 bootstrap samples).

| Model | MSE | R<sup>2</sup> | MAE | Rank Accuracy |  |  |
| --- | --- | --- | --- | --- | --- | --- |
|  |  |  |  | Lower | Middle | Upper |
| MLR | 0.035 | 0.155 | 0.142 | 0.570 | 0.403 | 0.601 |
|  | (0.031-0.149) | (0.000-0.179) | (0.127-0.150) | (0.516-0.628) | (0.372-0.433) | (0.537-0.665) |
| Random Forest Regression | 0.022 | 0.447 | 0.117 | 0.598 | 0.404 | 0.639 |
|  | (0.021-0.024) | (0.400-0.482) | (0.114-0.121) | (0.569-0.625) | (0.378-0.430) | (0.610-0.665) |
| XGBoost Regression | **0.021** | **0.482** | **0.112** | **0.609** | **0.413** | **0.650** |
|  | **(0.019-0.023)** | **(0.442-0.516)** | **(0.109-0.116)** | **(0.582-0.633)** | **(0.387-0.438)** | **(0.624-0.675)** |

Feature importance determined by SHAP analysis ranked the Pre-breakfast tag as being the most important, followed by Post-breakfast, Post-dinner, and Post-lunch tags. This was similar to the MLR coefficients which ranked Pre-breakfast as the most important, followed by Post-dinner, Post-breakfast, and Post-lunch tags. Results for the feature ranking of different Tags can be found in Supplementary Figure C1 and Table C2.

Non-linear, tree-based ensemble regression models achieved higher performance for this task, compared to an MLR model. Thus, as the XGBoost model out-performed both Random Forest and MLR models, especially with respect to the R2 value, we chose to use XGBoost for the development of all subsequent models.

Overall, performance across different models, did not differ substantially from the baseline model trained on only Tags (MSE, R2, MAE of 0.021 [CI 0.019-0.023], 0.482 [0.442-0.516], and 0.112 [0.109-0.116], respectively), upon the sequential addition of feature sets (MSE and MAE were 0.021 [CIs range 0.019-0.023] and 0.112 [CIs range 0.108-0.116], respectively, across all models). However, models that included High-Readings, performed slightly better (mean R2 of 0.486 [CIs range 0.442-0.521]), with the model trained on Tags and High-Readings achieving the best performance (R2 0.488 [CI 0.446-0.521]). Similar results were achieved with respect to rank accuracy, as lower-, middle-, and upper-bounds increased in accuracy with the addition of High-Readings (lower-, middle-, and upper-bound mean accuracies of 0.614 [CIs range 0.586-0.643], 0.417 [0.391-0.444], 0.655 [0.628-0.681]). Again, the model using Tags and High-Readings achieved the highest rank accuracies (lower-, middle-, and upper-bound accuracies of 0.616 [CI 0.589-0.643], 0.418 [0.394-0.444], 0.655 [0.628-0.681], respectively), followed closely by the model trained on Tags, High-Readings, and EHR (lower-, middle-, and upper-bound accuracies of 0.615 [CI 0.589-0.641], 0.418 [0.392-0.442], 0.655 [0.628-0.681], respectively). However, when comparing the outputs of the different models, they do not significantly differ from the model which is trained solely on Tags (model trained on Tags and High-Readings, p = 0.309 using the Wilcoxon Signed Rank Test; model trained on Tags, High-Readings, and EHR, p = 0.232). This suggests that blood glucose measurements themselves are collectively the most influential features for determining impending blood glucose anomalies. This is further confirmed by SHAP analysis, where Tags and High-Readings are consistently ranked highest in terms of feature importance across all models (SHAP results can be found in Appendix C of the Supplementary Material). The addition of Gradients, EHR, or the combination of both, did not appear to improve model performance over the corresponding models without these feature sets (p > 0.05). A full list of p-values comparing models to the baseline can be found in Supplementary Table C3.

As there were many missing values present for BMI in our dataset, we did not include it in our main analyses. However, we did perform preliminary analyses, including BMI as a feature in model development (using a subsequently reduced dataset). Additionally, we also performed analyses using the same dataset filtered to only include samples with blood glucose measurements between [1, 31]. Results for both can be found in Appendix D of the Supplementary Material.

### 4.2. External Validation

To demonstrate the generalizability of our method, we performed external validation on a cohort of women from RBH. The distribution of the predictor variable (Figure. 4b) is similar to that of the training set used during model development (Figure. 3).

**Figure 4:**
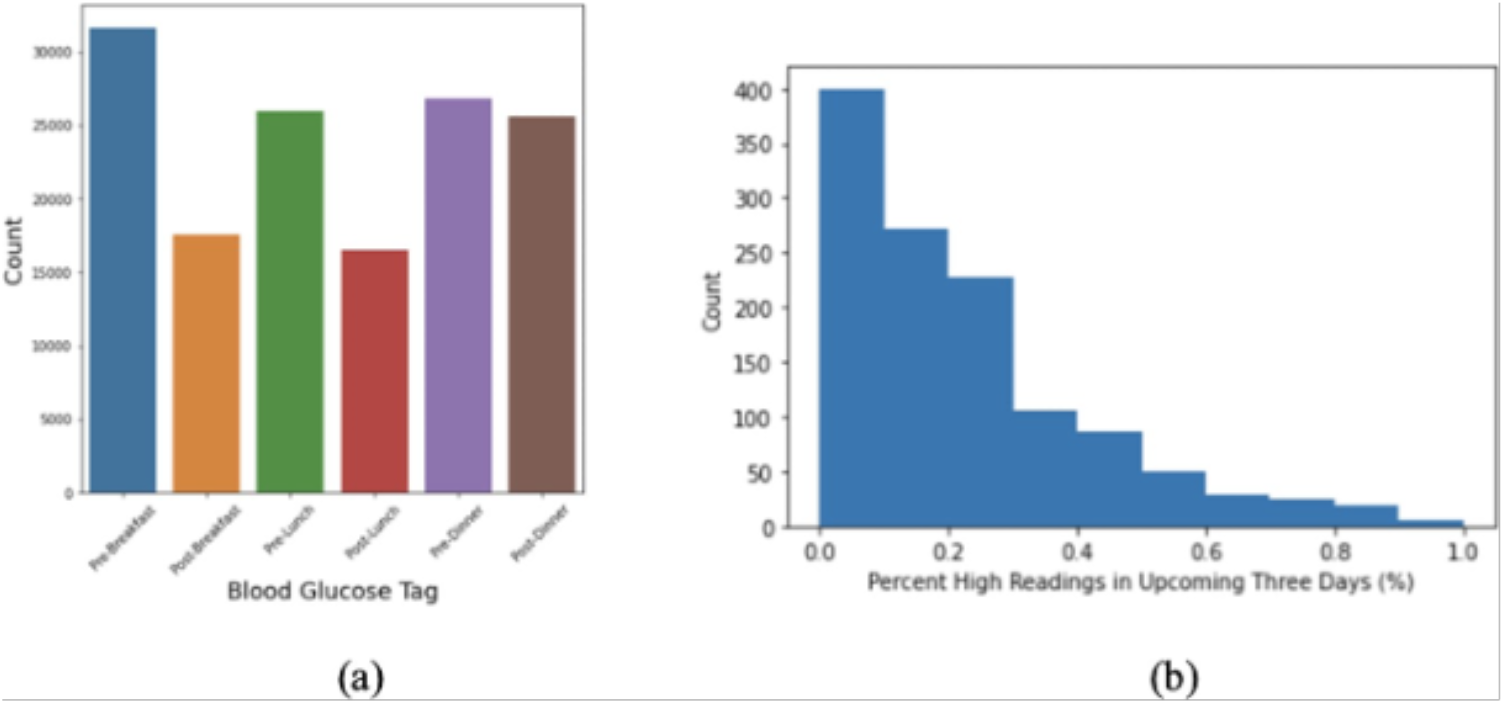
RBH dataset (a) Blood glucose readings for each tag across all pregnancies; (b) Percentage of high-reading alerts in the three-day prediction windows.

As the model using Tags and High-Readings achieved the best performance compared to other model variations, we used this as the model for external validation. We also compared this to the baseline model trained solely on Tags, as we found that they were not significantly different (p>0.05).

As shown in Table 5, when applied to the RBH cohort, our models achieved similar scores to those previously achieved from internal validation. The model trained on Tags achieved MSE, R2, and MAE scores of 0.021 (CI 0.020-0.021), 0.507 (0.494-0.519), 0.110 (0.109-0.111), respectively (compared to internal validation scores for MSE, R2, and MAE of 0.021 [CI 0.019-0.023], 0.482 [0.442-0.516], and 0.112 [0.109-0.116], respectively). The model trained on Tags and High-Readings achieved MSE, R2, and MAE scores of 0.020 (CI 0.020-0.021), 0.519 (0.505-0.530), 0.108 (0.107-0.110), respectively (compared to internal validation scores for MSE, R2, and MAE of 0.021 [CI 0.019-0.022], 0.488 [0.446-0.521], and 0.112 [0.108-0.116], respectively). The similarity in scores suggests that our model was not overfitted to the training set, and thus, is generalizable across external cohorts of patients. Additionally, as previously shown, the addition of High-Readings achieved better performance, overall, than the baseline model without this feature (Wilcoxon Signed Rank Test, p = 0.004).

## 5. Discussion

In this study, we developed a data-driven machine learning model to identify patients at risk of exhibiting high blood glucose levels (hyperglycaemia). This is a crucial task, as there is an appreciation that there exists a spectrum of disease and outcomes, and assigning all women to the same care pathway is not patient-centered and does not necessarily provide the best care for each woman. Furthermore, with increasing prevalence of GDM and limited healthcare resources, it is important for healthcare providers to tailor care delivery to women who need it – providing a proportionate response to care delivery depending on glycaemic control and other risk factors.

This study demonstrates the first machine learning-based stratification system for quantifying hyperglycaemia risk in GDM clinics, and is not limited to the existing GDm-Health platform. The model presented can be used by any GDM clinic if they have access to patients’ daily blood glucose data, and such a tool could be used to identify patients who require more urgent clinical review or need an adjustment to their current treatment. In order to translate this tool into clinical practice, future studies can consider converting the model results into a risk score (e.g. thresholding the regression scores into categories, combining regression scores with other clinical features to define the degree of risk).

We found that tree-based ensemble models significantly outperformed a linear model. This may be due to their inherent ability to consider non-linear effects of the features. Additionally, tree-based models are less sensitive extreme values (e.g. outliers, any data measurement errors which can occur from the data collection process) compared to linear regression. This is further demonstrated by MLR performing better on the filtered dataset, where extreme blood glucose values and sensor errors were removed prior to training (Supplementary Table D3).

There are several limitations to this study. Firstly, the MSE, R2, MAE, and rank accuracy scores suggest that this model can perform moderately accurately for predicting high blood glucose readings, especially those in the upper-bound group. However, the scores achieved require improvement in order to be suitable for clinical practice. This lower performance may be due to the small dataset size used in training, data imbalance, and possible clinical confounders that were not considered during model development. Further development needs to be done, with clinical experts, to both increase the sample size and determine what confounders should be included in the study. In terms of the algorithmic approach, a weighted regression model may help improve model fitting.

Additionally, different window sizes can be considered since it may be necessary to preserve a higher degree of granularity; binning may not be accurate enough to track and evaluate blood glucose patterns and is heavily biased on the sample population.

As blood glucose measurements are self-measured, there is variation in the number and length of recordings completed by each patient. Thus, when splitting the data into windows, there was an unequal number of samples contributed from different patients. Ideally, the number of samples available from each patient is balanced; however, in the real world this difficult to achieve, as women may be diagnosed at different points in their pregnancy and may have very different lifestyles, making it difficult to collect consistent measurements across all patients. If enough data is available, one possible solution can be regularizing the number of samples contributed by each patient (reducing the bias contributed from any individual patient). Similarly, future models can collectively consider multiple windows per patient rather than treating each window separately.

The specification of when a patient started or stopped taking GDM-related medication was not clear in our data. We considered women who had taken any medication for blood glucose control (metformin or insulin) as the medication group and the remainder as the non-medication group. Patients who changed from non-medication to medication during the data collection period of the study, were considered as part of the medication group. This is a limitation of this study, as it may have impacted the models’ ability to confidently differentiate different groups and accurately predict the high-reading percentages. By comparing model performance results (Table 4) and SHAP values (Supplementary Figures C1-C8), we found that including the medication feature did not significantly change our results when compared to the baseline model. Thus, future analyses should be performed, with more data, to confirm the effectiveness of including this feature.

**Table 4:** Overall performance and rank accuracy of XGBoost models trained on different feature set combinations. Results are reported alongside 95% confidence intervals (calculated using 1000 bootstrap samples).

| Feature Sets<br>Used | MSE | R<sup>2</sup> | MAE | Rank Accuracy |  |  |
| --- | --- | --- | --- | --- | --- | --- |
|  |  |  |  | Lower | Middle | Upper |
| **Tags** | 0.021<br>(0.019-0.023) | 0.482<br>(0.442-0.516) | 0.112<br>(0.109-0.116) | 0.609 (0.582-0.633) | 0.413 (0.387-0.438) | 0.650<br>(0.624-0.675) |
| **Tags,**<br>**Gradients** | 0.021<br>(0.019-0.023) | 0.480<br>(0.442-0.517) | 0.112<br>(0.108-0.116) | 0.608 (0.580-0.635) | 0.412 (0.385-0.437) | 0.650<br>(0.626-0.675) |
| **Tags, High-**<br>**Readings** | 0.021<br>(0.019-0.022) | **0.488**<br>**(0.446-**<br>**0.521)** | 0.112<br>(0.108-0.116) | **0.616 (0.589-**<br>**0.643)** | **0.418 (0.394-**<br>**0.444)** | **0.655**<br>**(0.628-**<br>**0.681)** |
| **Tags,**<br>**Gradients,**<br>**High-**<br>**Readings** | 0.021<br>(0.019-0.022) | 0.484<br>(0.444-0.515) | 0.112<br>(0.108-0.115) | 0.611 (0.586-0.638) | 0.416 (0.391-0.441) | 0.654<br>(0.628-0.678) |
| **Tags, EHR** | 0.021<br>(0.019-0.023) | 0.480<br>(0.440-0.510) | 0.112<br>(0.108-0.116) | 0.611 (0.583-0.635) | 0.412 (0.387-0.438) | 0.650<br>(0.622-0.676) |
| **Tags, EHR,**<br>**Gradients** | 0.021<br>(0.019-0.023) | 0.480<br>(0.437-0.514) | 0.112<br>(0.108-0.116) | 0.610 (0.581-0.635) | 0.414 (0.387-0.440) | 0.650<br>(0.623-0.676) |
| **Tags, EHR,**<br>**High-**<br>**Readings** | 0.021<br>(0.019-0.022) | 0.486<br>(0.448-0.518) | 0.112<br>(0.108-0.116) | 0.615 (0.589-0.641) | **0.418 (0.392-**<br>**0.442)** | **0.655**<br>**(0.628-**<br>**0.681)** |
| **Tags, EHR,**<br>**Gradients,**<br>**High-**<br>**Readings** | 0.021<br>(0.019-0.023) | 0.484<br>(0.442-0.514) | 0.112<br>(0.108-0.116) | 0.613 (0.586-0.643) | 0.417 (0.392-0.444) | 0.654<br>(0.629-0.680) |

**Table 5:** Overall performance and rank accuracy of XGBoost models externally validated on RBH cohort. Results are reported alongside 95% confidence intervals (calculated using 1000 bootstrap samples).

| Feature Sets Used | MSE | R<sup>2</sup> | MAE | Rank Accuracy |  |  |
| --- | --- | --- | --- | --- | --- | --- |
|  |  |  |  | Lower | Middle | Upper |
| Tags | 0.021<br>(0.020-0.021) | 0.507<br>(0.494-0.519) | 0.110<br>(0.109-0.111) | 0.624 (0.609-0.639) | 0.413 (0.391-0.430) | **0.627 (0.612-0.639)** |
| Tags, High-Readings | **0.020 (0.020-0.021)** | **0.519 (0.505-0.530)** | **0.108 (0.107-0.110)** | **0.639 (0.624-0.654)** | **0.420 (0.398-0.440)** | 0.624 (0.607-0.639) |

Data missingness is another limitation, as we used real-world clinical data sets. After we removed samples with values missing values from any of our selected features, the size of data available for training and testing was reduced to 70% and 25% of the original dataset size in the OUH and RBH datasets, respectively. Thus, future analyses would greatly benefit from more data or the application of different data imputation methods. For this study, we did not have enough data to impute missing values in a way that would be biologically accurate.

In general, overall performance did not differ significantly with the addition of non-blood-glucose features. This may be due to the size of our dataset, as a larger number of features often require a larger sample size for training. Additionally, it is difficult to understand how different behavioural, physiological, and genetic factors independently and collectively affect GDM, and thus, there may be other factors that are better suited to use in the model than the ones we tested. For example, known diabetes risk factors (e.g. high BMI, previously having a macrosomic baby, being from an ethnicity with a higher prevalence of GDM), may be important in blood glucose prediction. In this paper, whilst we were not able to test these features, this reflects the reality of features directly accessible by most GDM clinics. Although BMI was not included in our main model development (due to many missing values), it was ranked moderately high in terms of feature importance in our preliminary analyses (Supplementary Table D2). Thus, future models can consider additional EHR features during development, as it may help improve model performance.

## 6. Conclusion

With the massive growth of digital sensors and electronic data continuing to saturate healthcare, machine learning will greatly support clinicians in optimizing healthcare utilization and facilitating patient care. This paper presents one of the largest clinical machine learning studies on GDM patient stratification and provides a proof-of-concept demonstration of how personalized patient care can be implemented for GDM patients. As there is currently no mechanism in place to predict those women at risk of hyperglycaemia, our study outlines and demonstrates a straightforward method for implementing proportionate care delivery based on features already available in many GDM clinics. Additionally, our framework has the potential to be extended to and used with many other predictor features and applications. Overall, machine learning in GDM is still a relatively new area; thus, additional model training and external validation is necessary to improve our understanding of GDM, clinical management, and ultimately, overall maternal and fetal health and care.

## Data Availability

Patient data cannot be shared due to data privacy.

## Author Contributions

Conceptualization, H.L., J.Y., J.H., and L.M.; methodology, J.Y. and H.L.; software, J.Y., and H.L.; validation, J.Y., and H.L.; formal analysis, J.Y.; investigation, J.Y., H.L and L.M.; resources, H.L., L.M; data curation, J.Y.; writing—original draft preparation, J.Y.; writing—review and editing, J.Y., H.L., J.H., L.M..; visualization, J.Y and H.L..; supervision, H.L. and L.M.; project administration, H.L.; funding acquisition, H.L. and D.C. All authors have read and agreed to the published version of the manuscript.

## Institutional Review Board Statement

This project is a part of the clinical study “Predictive monitoring and management of pregnant women with gestational diabetes mellitus”. The study was conducted in accordance with the Declaration of Helsinki, and approved by the UK Health Research Authority (REC reference: 21/HRA/3733 and date of 1 August 2020.

## Informed Consent Statement

Informed consent was obtained from all subjects involved in the study.

## Data Availability Statement

Patient data cannot be shared due to data privacy.

## Acknowledgments

We express our sincere thanks to all patients and staff across Oxford University Hospitals NHS Foundation Trust and Royal Berkshire Foundation Trust. We additionally express our gratitude to Professor Lionel Tarassenko and Dr. Carmelo Verlardo for their work in the GDm-Health and support in the RBH data extraction.

## References

1. IDF Atlas, The International Diabetes Federation Altas Tenth edition (2021). https://idf.org/aboutdiabetes/what-is-diabetes/facts-figures.html.

2. American Diabetes Association. (2003). Report of the expert committee on the diagnosis and classification of diabetes mellitus. Diabetes care, 26(suppl 1), s5–s20.

3. Oskovi-Kaplan, Z. A., Ozgu-Erdinc, A. S. (2020). Management of gestational diabetes mellitus. In Diabetes: from Research to Clinical Practice (pp. 257–272). Springer, Cham.

4. Martis, R., Crowther, C. A., Shepherd, E., Alsweiler, J., Downie, M. R., Brown, J. (2018). Treatments for women with gestational diabetes mellitus: an overview of Cochrane systematic reviews. Cochrane Database of Systematic Reviews, (8).

5. Turok, D. K., Ratcliffe, S., Baxley, E. G. (2003). Management of gestational diabetes mellitus. American family physician, 68(9), 1767–1772.

6. Mackillop, L., Loerup, L., Bartlett, K., Farmer, A., Gibson, O. J., Hirst, J. E., Tarassenko, L. (2014). Development of a real-time smartphone solution for the management of women with or at high risk of gestational diabetes. Journal of diabetes science and technology, 8(6), 1105–1114.

7. National Institute for Health and Care Excellence. (2015, Feb 15) Diabetes in pregnancy: Management of diabetes and its complications from preconception to the postnatal period. NG3. https://www.nice.org.uk/guidance/ng3.

8. Li, Y., Ren, X., He, L., Li, J., Zhang, S., & Chen, W. (2020). Maternal age and the risk of gestational diabetes mellitus: a systematic review and meta-analysis of over 120 million participants. Diabetes research and clinical practice, 162, 108044.

9. Bochkur Dratver, M. A., Arenas, J., Thaweethai, T., Yu, C., James, K., Rosenberg, E. A.,Powe, C. E. (2021). Longitudinal changes in glucose during pregnancy in women with gestational diabetes risk factors. Diabetologia, 1–11.

10. Finneran, M. M., & Landon, M. B. (2018). Oral agents for the treatment of gestational diabetes. Current diabetes reports, 18(11), 1–8.

