## Supplementary Material for "Machine Learning-Based Risk Stratification for Gestational Diabetes Management"

**Jenny Yang et al., 2022**

**Appendix A: Software packages and Implementation**

Models were implemented using Python (v3.6.9). Scikit Learn (v0.24.1) was used for implementing multi linear regression and calculating performance metrics. XGBoost models were implemented using the XGBoost library (v1.3.3).

**Appendix B: Summary Characteristics of Processed Datasets**

**1.** ***Oxford University Hospitals NHS Foundation Trust (OUH)***:

**Supplementary Table B1**: OUH summary statistics for features used in model development. Population mean presented alongside standard deviation (SD) and 25th-, 50th-, and 75th-percentiles.

| Feature: | Count | Mean | SD | 25% | 50% | 75% |
|---|---|---|---|---|---|---|
| **Pre-Breakfast Tag** | 840 | 5.335 | 1.531 | 4.8 | 5.200 | 5.648 |
| **Post-Breakfast Tag** | 840 | 7.066 | 2.712 | 6.05 | 6.767 | 7.588 |
| **Post-Lunch Tag** | 840 | 6.664 | 2.185 | 5.881 | 6.467 | 7.150 |
| **Post-Dinner Tag** | 840 | 6.814 | 2.058 | 5.968 | 6.664 | 7.350 |
| **High-Readings Proportion** | 840 | 0.249 | 0.225 | 0.083 | 0.222 | 0.375 |
| **Pre-Breakfast Gradient** | 840 | -0.015 | 0.636 | -0.25 | 0.000 | 0.200 |
| **Post-Breakfast Gradient** | 840 | -0.066 | 1.485 | -0.551 | -0.050 | 0.500 |
| **Post-Lunch Gradient** | 840 | 0.019 | 1.024 | -0.550 | 0.000 | 0.550 |
| **Post-Dinner Gradient** | 840 | -0.044 | 0.994 | -0.550 | 0.000 | 0.462 |
| **Gestational Age** | 840 | 221.830 | 39.966 | 203.083 | 229 | 251.5 |
| **Medication** | 840 | 0.356 | 0.479 | 0 | 0 | 1 |
| **BMI** | 452 | 30.964 | 6.799 | 25.910 | 30.425 | 34.963 |

**2.** ***Royal Berkshire Hospitals NHS Foundation Trust (RBH)***:

**Supplementary Table B2**: RBH summary statistics for features used in model development. Population mean presented alongside standard deviation (SD) and 25th-, 50th-, and 75th-percentiles.

| Feature: | Count | Mean | SD | 25% | 50% | 75% |
|---|---|---|---|---|---|---|
| **Pre-Breakfast Tag** | 186 | 5.155 | 0.889 | 4.6 | 5.028 | 5.425 |
| **Post-Breakfast Tag** | 186 | 6.84 | 1.204 | 6.037 | 6.658 | 7.4 |
| **Post-Lunch Tag** | 186 | 9.88 | 44.07 | 5.969 | 6.6 | 7.347 |
| **Post-Dinner Tag** | 186 | 6.86 | 1.164 | 6.153 | 6.709 | 7.296 |
| **High-Readings Proportion** | 186 | 0.207 | 0.187 | 0.067 | 0.167 | 0.3 |

**Appendix C: Model Development**

1. *Final Hyperparameter Values Used for Models:*

**Supplementary Table C1**: Hyperparameter values used in model development (XGBoost)

| Feature set | Colsample (tree) | Gamma | Learning rate | Max depth | N estimators | Sub-sample |
|---|---|---|---|---|---|---|
| **Tags** | 0.5 | 0.9 | 0.05 | 3 | 58 | 0.7 |
| **Tags, Gradients** | 0.7 | 0.9 | 0.05 | 3 | 57 | 0.9 |
| **Tags, High-Readings** | 0.5 | 0.9 | 0.05 | 3 | 59 | 0.9 |
| **Tags, Gradients, High-Readings** | 0.7 | 0.5 | 0.05 | 3 | 58 | 0.9 |
| **Tags, EHR** | 0.5 | 0.9 | 0.05 | 3 | 59 | 0.9 |
| **Tags, EHR, Gradients** | 0.7 | 0.9 | 0.05 | 3 | 59 | 0.9 |
| **Tags, EHR, High-Readings** | 0.5 | 0.5 | 0.05 | 3 | 46 | 0.9 |
| **Tags, EHR, Gradients, High-Readings** | 0.7 | 0.5 | 0.05 | 3 | 47 | 0.9 |

**Supplementary Table C2**: Linear Regression Coefficients for model trained on Tags.

| Before Breakfast | After Breakfast | After Lunch | After Dinner |
|---|---|---|---|
| **0.11896724** | 0.02644718 | 0.0061178 | 0.05830037 |

2. *SHAP Analysis Results:*

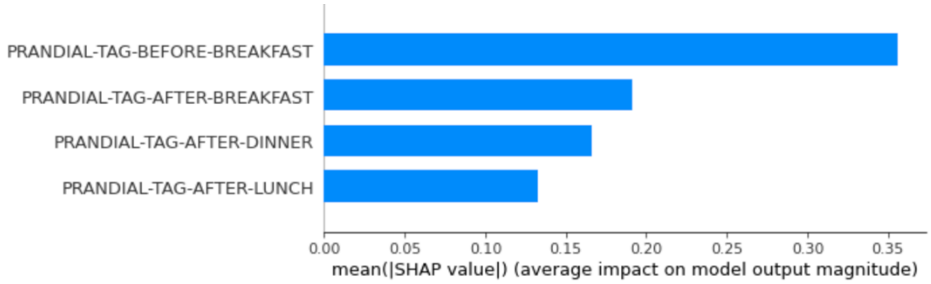


**Supplementary Figure C1**: SHAP feature importance ranking for model trained on Tags.

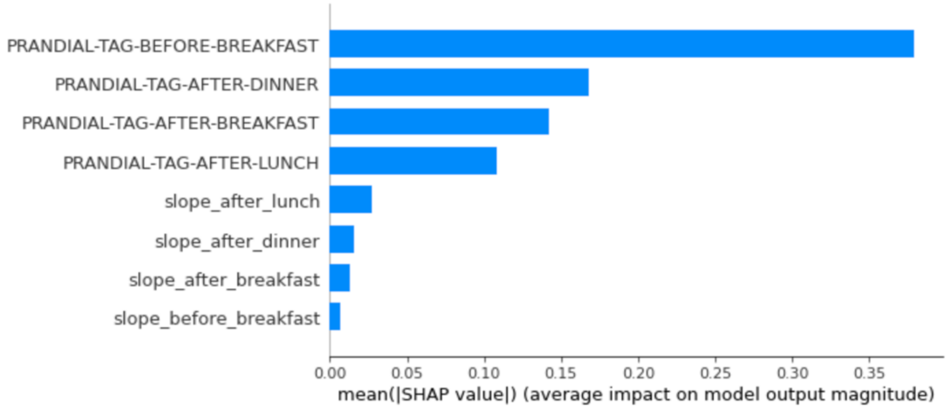


**Supplementary Figure C2**: SHAP feature importance ranking for model trained on Tags and Gradients.

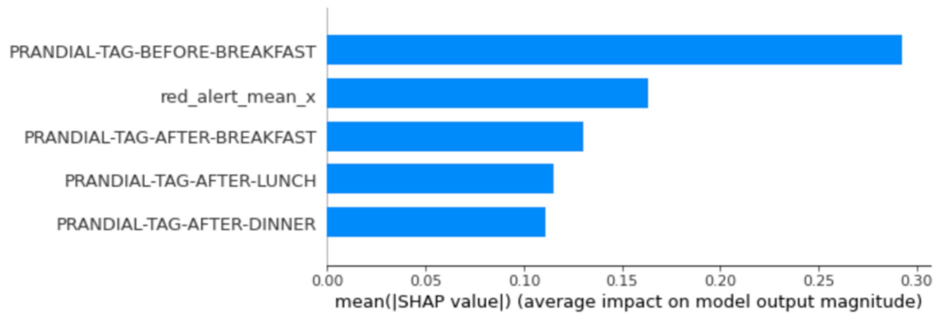


**Supplementary Figure C3**: SHAP feature importance ranking for model trained on Tags and High-Readings.

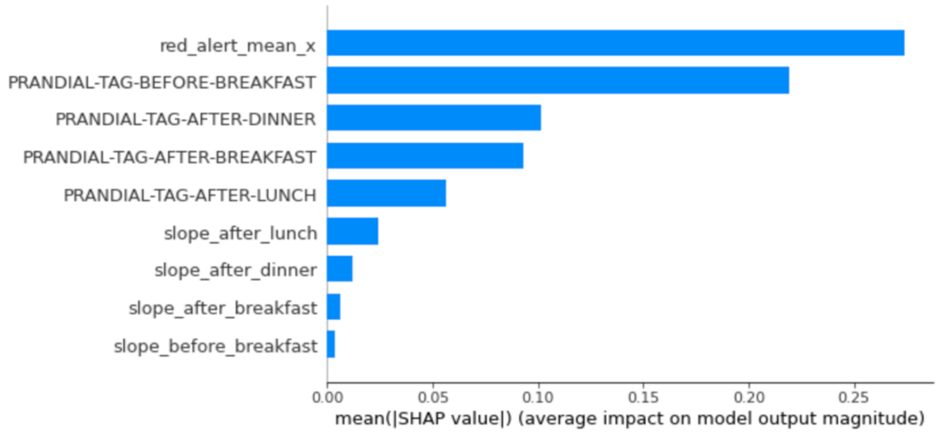


**Supplementary Figure C4**: SHAP feature importance ranking for model trained on Tags, Gradients, and High-Readings.

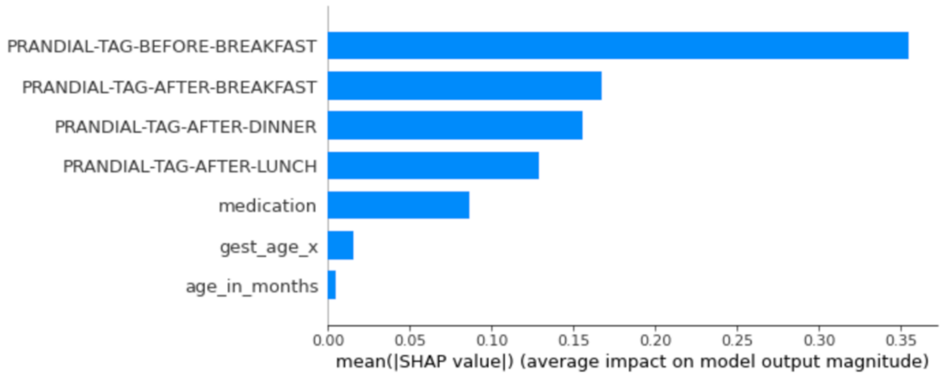


**Supplementary Figure C5**: SHAP feature importance ranking for model trained on Tags and EHR features.

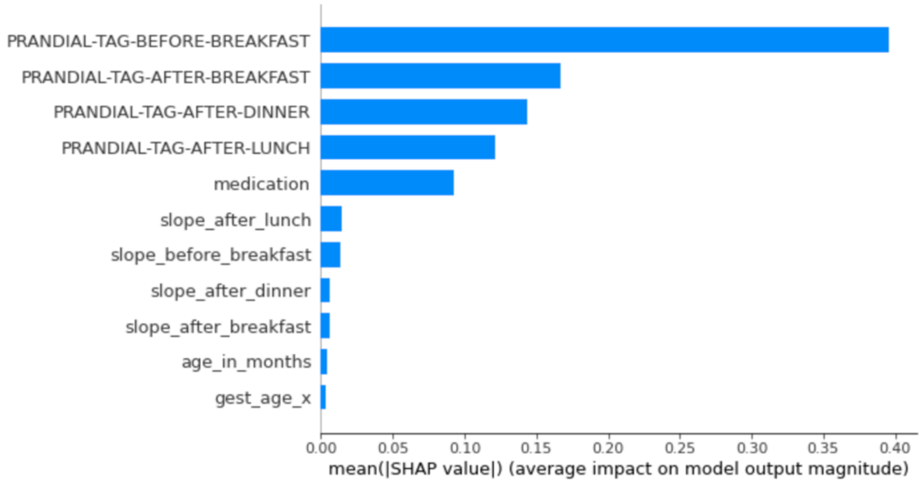


**Supplementary Figure C6**: SHAP feature importance ranking for model trained on Tags, Gradients, and EHR features.

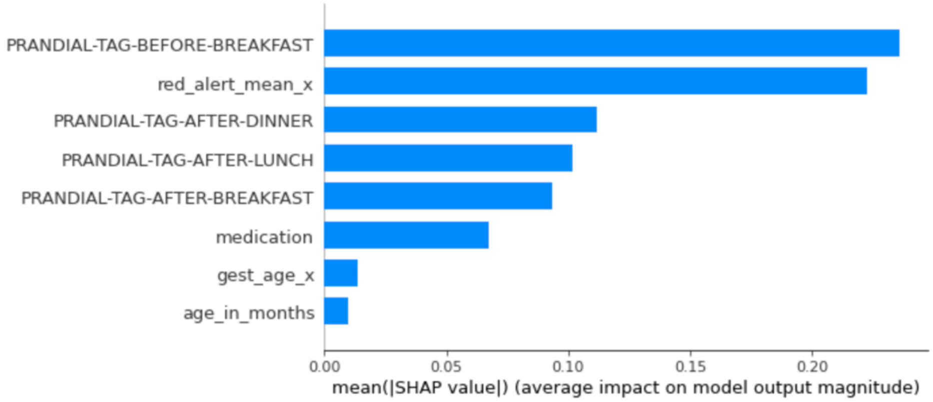


**Supplementary Figure C7**: SHAP feature importance ranking for model trained on Tags, High-Readings, and EHR features.

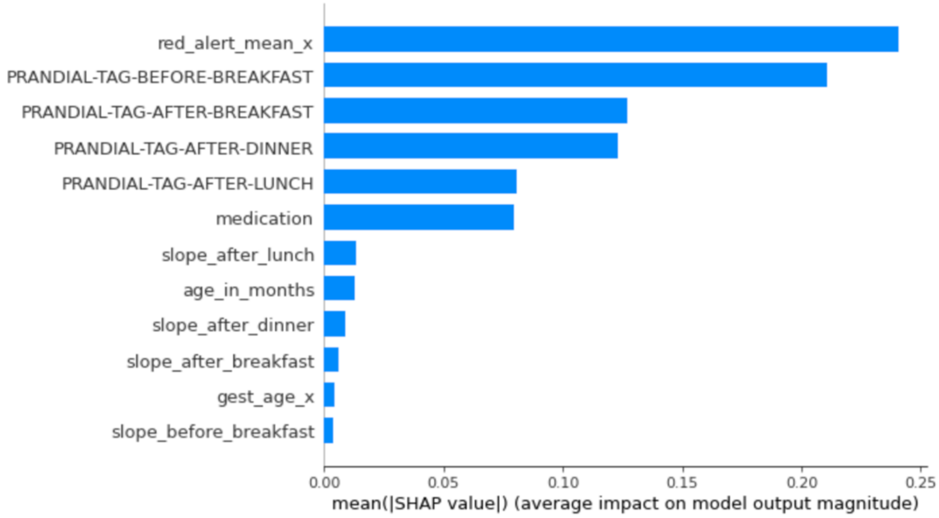


**Supplementary Figure C8**: SHAP feature importance ranking for model trained on Tags, Gradients, High-Readings, and EHR features.

**Supplementary Table C3**: p-values (Wilcoxon Signed Rank Test) comparing outputs of each model (trained on different feature set combinations) to the baseline model (trained solely on Tags). Wilcoxon Signed Rank Test implemented using the statistics package from the SciPy library (scipy.stats).

| **Feature Sets Used** | **p-value** |
|---|---|
| Tags, Gradients | <0.0001 |
| Tags, High-Readings | 0.309 |
| Tags, Gradients, High-Readings | 0.308 |
| Tags, EHR | <0.0001 |
| Tags, EHR, Gradients | <0.0001 |
| Tags, EHR, High-Readings | 0.232 |
| Tags, EHR, Gradients, High-Readings | 0.124 |

**Appendix D: Additional Analyses**

1. ***Final Hyperparameter Values Used for Models***:

**Supplementary Table D1**: Hyperparameter values used in model development (XGBoost) for additional experiments: 1) Including BMI as a feature, 2) Using a dataset filtered for blood glucose range [1,30]

| **Feature set** | **Colsample (tree)** | **Gamma** | **Learning rate** | **Max depth** | **N estimators** | **Sub-sample** |
|---|---|---|---|---|---|---|
| **Tags, Gradients, High-readings, EHR (including BMI)** | 0.5 | 0.9 | 0.05 | 3 | 51 | 0.9 |
| **Tags** | 0.5 | 0.7 | 0.05 | 3 | 59 | 0.9 |

2. ***Model Development on a Training Set Including BMI as a Predictor Variable***:

**Supplementary Table D2**: Results of models trained on all features, including BMI, using the reduced dataset (362 patients in training set, corresponding to 2,500 windows in test set; 90 patients in testing, corresponding to 601 windows).

| **Feature Sets Used** | **Model** | **MSE** | **$R^2$** | **MAE** | **Rank Accuracy** | | |
|---|---|---|---|---|---|---|---|
| | | | | | **Lower** | **Middle** | **Upper** |
| **Tags, Gradients, High-readings, EHR (including BMI)** | MLR | 0.023 (0.020-0.113) | 0.404 (0.000-0.506) | 0.116 (0.109-0.129) | 0.624 (0.588-0.660) | 0.433 (0.396-0.466) | 0.642 (0.605-0.679) |
| | Random Forest Regression | 0.022 (0.020-0.024) | 0.475 (0.406-0.527) | 0.114 (0.109-0.119) | 0.615 (0.576-0.655) | 0.426 (0.393-0.459) | 0.644 (0.604-0.679) |
| | XGBoost Regression | 0.022 (0.020-0.025) | 0.470 (0.410-0.513) | 0.113 (0.107-0.119) | 0.612 (0.575-0.654) | 0.421 (0.385-0.455) | 0.643 (0.607-0.679) |

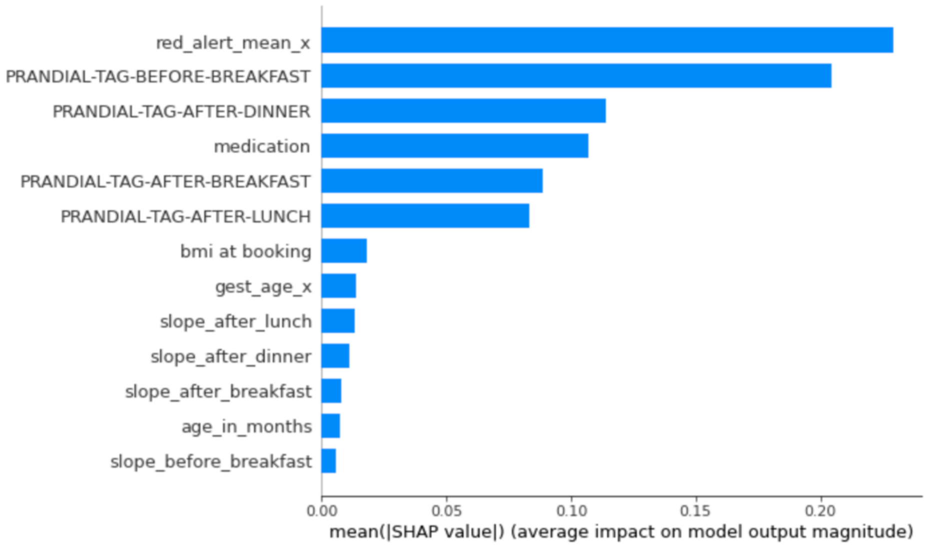


**Supplementary Figure D1**: SHAP ranking for all features, including BMI, using reduced dataset (362 patients in training set, corresponding to 2,500 windows in test set; 90 patients in testing, corresponding to 601 windows).

**3.** ***Model Development on a Training Set with a Fixed Blood Glucose Range***:

**Supplementary Table D3**: Results of model trained on Using a filtered Dataset including only blood glucose levels [1, 31] (656 patients in training set, corresponding to 4,580 windows; 163 patients in test set, corresponding to 1166 windows):

| **Feature Sets Used** | **Model** | **MSE** | **$R^2$** | **MAE** | **Rank Accuracy** | | |
|---|---|---|---|---|---|---|---|
| | | | | | **Lower** | **Middle** | **Upper** |
| **Tags, High-readings, Intervention, Gradients** | MLR | 0.022 (0.020-0.024) | 0.445 (0.397-0.481) | 0.114 (0.111-0.118) | 0.613 (0.587-0.639) | 0.418 (0.393-0.444) | 0.651 (0.621-0.678) |
| | Random Forest Regression | 0.022 (0.020-0.024) | 0.445 (0.399-0.479) | 0.116 (0.113-0.120) | 0.596 (0.567-0.624) | 0.403 (0.379-0.429) | 0.640 (0.610-0.668) |
| | XGBoost Regression | 0.020 (0.019-0.022) | 0.482 (0.437-0.514) | 0.111 (0.108-0.115) | 0.605 (0.580-0.632) | 0.412 (0.388-0.439) | 0.651 (0.625-0.678) |
